# Longitudinal patterns and determinants of statin adherence in over one million individuals from Finland and Italy

**DOI:** 10.64898/2026.01.26.26344722

**Authors:** Andrea Corbetta, Katherine M. Logan, Matteo Ferro, Markus Perola, Andrea Ganna, Emanuele Di Angelantionio, Francesca Ieva

## Abstract

Medication adherence is critical for effective management of chronic diseases and reducing healthcare burdens. Statins, commonly prescribed for cardiovascular disease prevention, require sustained, lifelong adherence, yet maintaining long-term adherence remains a significant challenge. Here, we analysed longitudinal electronic health records from over one million statin users in Finland and Italy to characterise adherence trajectories and their determinants. Using functional data analysis, we identified five distinct adherence patterns, with consistently high adherence being the most prevalent across both populations. Younger age, socioeconomic vulnerability, and statin use for primary prevention were consistently associated with a higher risk of declining adherence over time. Sex differences were observed in Italy but not in Finland, where divorced status and health-related educational background were also associated with declining adherence. Despite differences in healthcare systems, several determinants of adherence were consistent across countries. These findings highlight shared behavioural factors underlying long-term statin use and suggest that population-level interventions tailored to patient subgroups defined by adherence patterns may help support sustained medication adherence.

## Introduction

Adherence to prescribed medication is a cornerstone of effective treatment and plays a key role in improving patient outcomes^1^ and reducing unnecessary healthcare expenditures.^2,3^ Statins, among the most widely prescribed medications (for example, atorvastatin was the most common medication prescribed in the US in 2022^4^), are central to both the prevention and management of cardiovascular disease. Often used alongside antihypertensives, statins are prescribed to individuals at risk of, or already affected by, cardiovascular conditions. Their efficacy in lowering cholesterol levels is well-established through randomised clinical trials.^5–7^ However, achieving optimal cholesterol control in routine clinical practice remains challenging, primarily due to poor long-term adherence to treatment, which requires consistent, lifelong medication use.^8^

Despite growing interest, the determinants of poor medication adherence remain incompletely understood and inconsistent across studies.^9^ Existing evidence is limited by reliance on self-reported adherence data from small, non-representative samples^10^, as well as by a narrow temporal focus, with long-term adherence trajectories, particularly beyond several years, remaining largely unexplored.^11,12^ Most prior research also conceptualises adherence as a static metric, ^13–15^ typically using a binary measure or threshold-based conversions of measures such as medication possession ratio (MPR) or proportion of days covered (PDC). ^16^ Although some studies have examined longitudinal adherence using group-based trajectory models (GBTM),^17,18^ these approaches rely on discretised measures and may fail to capture the full complexity and temporal dynamics of medication-taking behaviour.

In this study, we analysed longitudinal electronic health records from two large-scale European cohorts: the entire population of long-term statin users in Finland (n = 544,878) and the Lombardy region of Italy (n = 613,478), totalling over 1.15 million individuals. We applied functional data analysis^19^ (FDA) to model adherence as a continuous, time-evolving behaviour, enabling a detailed characterisation of adherence trajectories and their temporal dynamics. Using this framework, we evaluated over a hundred demographic, socioeconomic, and clinical features in multivariable models to identify predictors associated with distinct adherence patterns (Figure 1).

**Figure 1:**
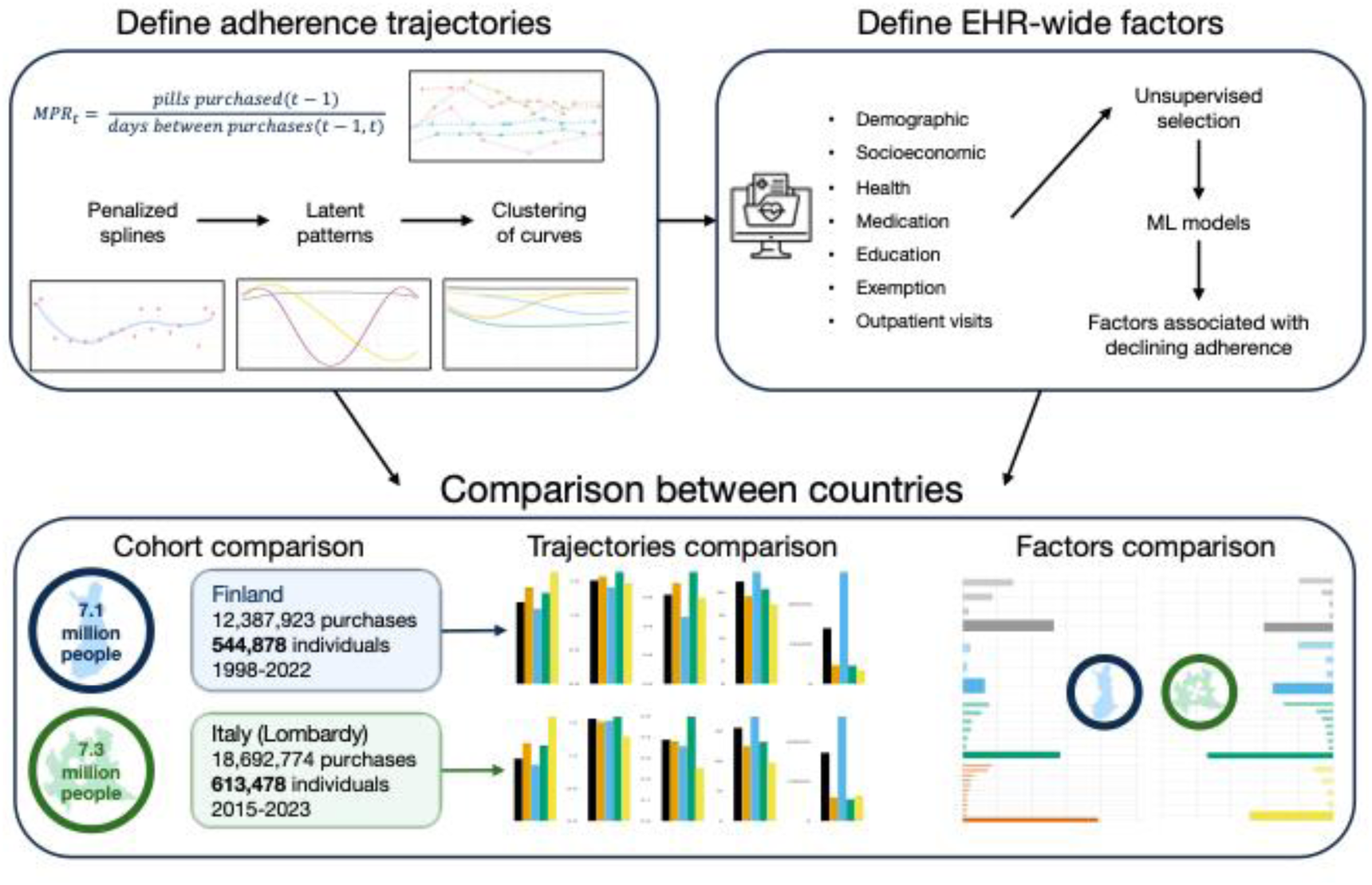
Graphical abstract Three steps of the analysis: definition of adherence trajectories, definition of baseline EHR-wide factors and modelling, comparison between countries

This study provides a comprehensive, longitudinal characterisation of statin adherence across two contrasting healthcare systems. By integrating advanced statistical modelling with high-resolution population-level data, we identify both shared and context-specific factors associated with declining adherence. These findings may inform precision-targeted interventions to support long-term medication adherence at scale.

## Results

### Adherence and prescribing patterns vary across countries

In Finland, 544,878 of 1,484,233 statin users between 1998 and 2017 met the inclusion criteria of at least five years of continuous treatment. The mean age was of 62 (SD 10), and 48% were female (Supplementary Data 1). Mean adherence, measured by MPR, was 0.88 (SD 0.10), with a median of 22 statin packages purchased and a median refill interval of 100 days (Figure 2a). Most individuals (279,749; 51%) remained on mid-potency statins throughout, 15% used low-potency, and 12.6% switched from low- to mid-potency. Only 9% had more than one potency switch (Table 1). In Italy, 613,478 of 1,861,547 statin users between 2012 and 2018 fulfilled the same criteria. Mean age was of 68 (SD 10) and 47% were female (Supplementary Data 1). Mean MPR was 0.82 (SD 0.12), with a median of 31 packages purchased and a median refill interval of 70 days (Figure 2a). Most individuals (66%) remained on mid-potency statins, 9% on high potency, and 7% switched from mild to high potency. Multiple potency switches were observed in 12% of individuals (Table 1). Age- and sex-specific prevalence of statin use was similar between countries, with slightly higher prevalence in Finland across most age groups (Spearman correlation = 0.91; Figure 2b).

**Figure 2:**
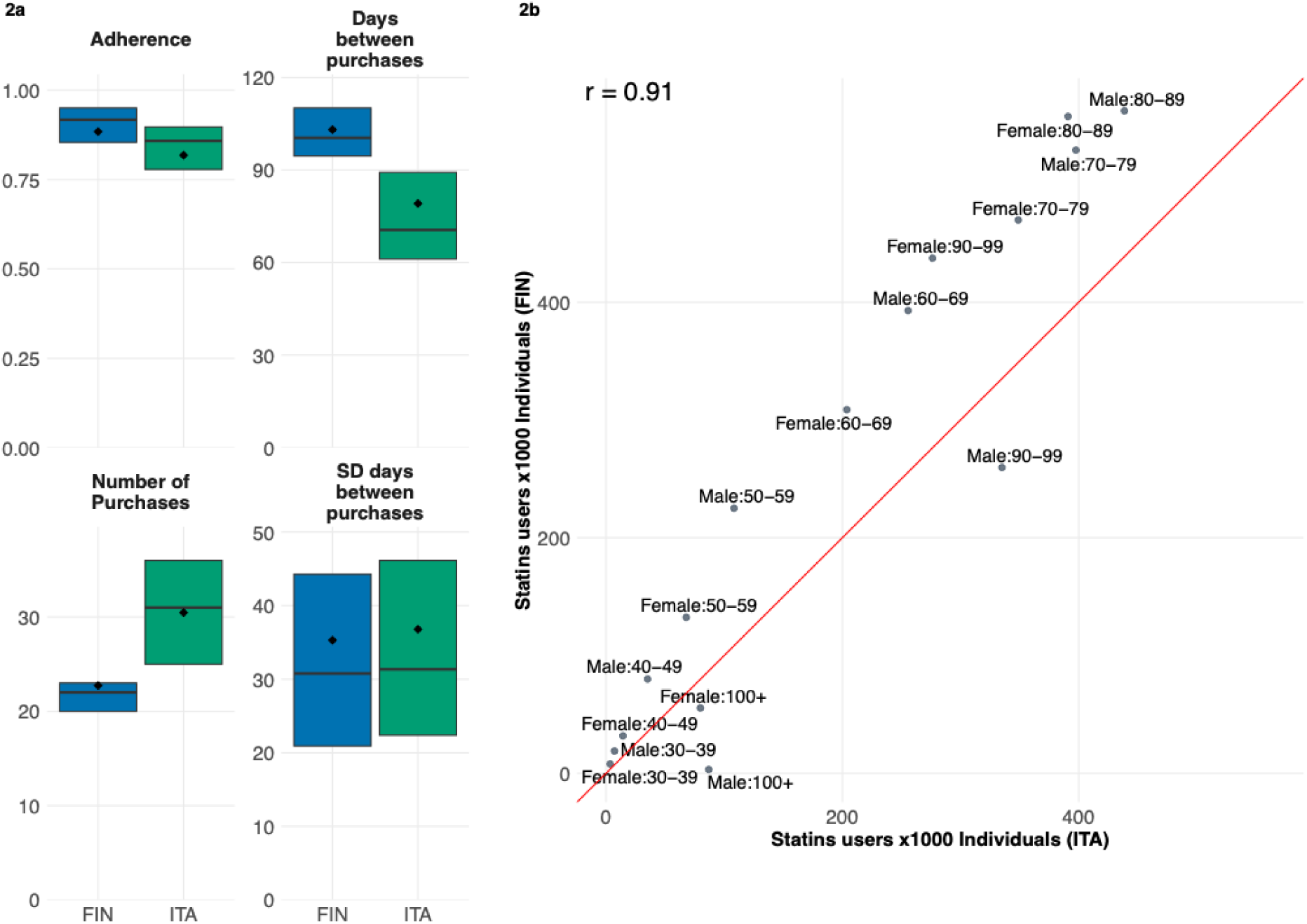
Adherence descriptives (a) Statins users per 1,000 individuals in Finland and Italy (Lombardy) by age and sex; (b) Faux boxplot of the mean MPR, mean days between purchase, number of purchases and SD of days between purchases

**Table 1:** Summary of potency switches Most prevalent sequences of statin potency switches (absolute and relative prevalence)

| Finland |  |  | Italy |  |  |
| --- | --- | --- | --- | --- | --- |
| Sequence | n | % | Sequence | n | % |
| Mid | 279,749 | 51.34% | Mid | 406,427 | 66.25% |
| Low | 86,242 | 15.83% | High | 54,990 | 8.96% |
| Low=Mid | 68,436 | 12.56% | Mid=High | 42,663 | 6.95% |
| Mid=High | 21,166 | 3.88% | High=Mid | 14,827 | 2.42% |
| Mid=Low | 15,412 | 2.83% | Mid=High=Mid | 13,653 | 2.23% |
| Mid=Low=Mid | 12,736 | 2.34% | Low | 9,483 | 1.55% |
| High | 9,167 | 1.68% | High=Mid=High | 7,312 | 1.19% |
| Low=Mid=Low=Mid | 6,154 | 1.13% | Mid=High=Mid=High | 7,204 | 1.17% |
| Low=Mid=Low | 4,942 | 0.91% | Mid==Mid | 6,919 | 1.13% |
| Mid=High=Mid | 4,747 | 0.87% | Low=Mid | 4,595 | 0.75% |

### Latent adherence trajectories captured by functional data analysis

Functional data analysis revealed common longitudinal adherence patterns in both cohorts. We modelled five-year sequences of MPR as smooth functions of time and applied functional principal component analysis^19^ (FPCA) to identify the main uncorrelated latent patterns in adherence trajectories (Figure 3a; Methods). In the Finnish cohort, the first principal component, reflecting consistent adherence, accounted for 57.9% of total variance. The second component (14.5%) captured increasing or decreasing trends, while the third (8.3%) represented mid-period peaks or drops, typically around 2.5 years. In the Italian cohort the first component explained 60.5% of the variance, followed by components representing monotonic trends (11.6%) and mid-period changes (7.0%), all holding the same interpretation as the Finnish cohort. (Supplementary Figure 2 and 3). Individual trajectories were summarised by FPCA scores, enabling a lower-dimensional representation of longitudinal adherence behaviour.

**Figure 3:**
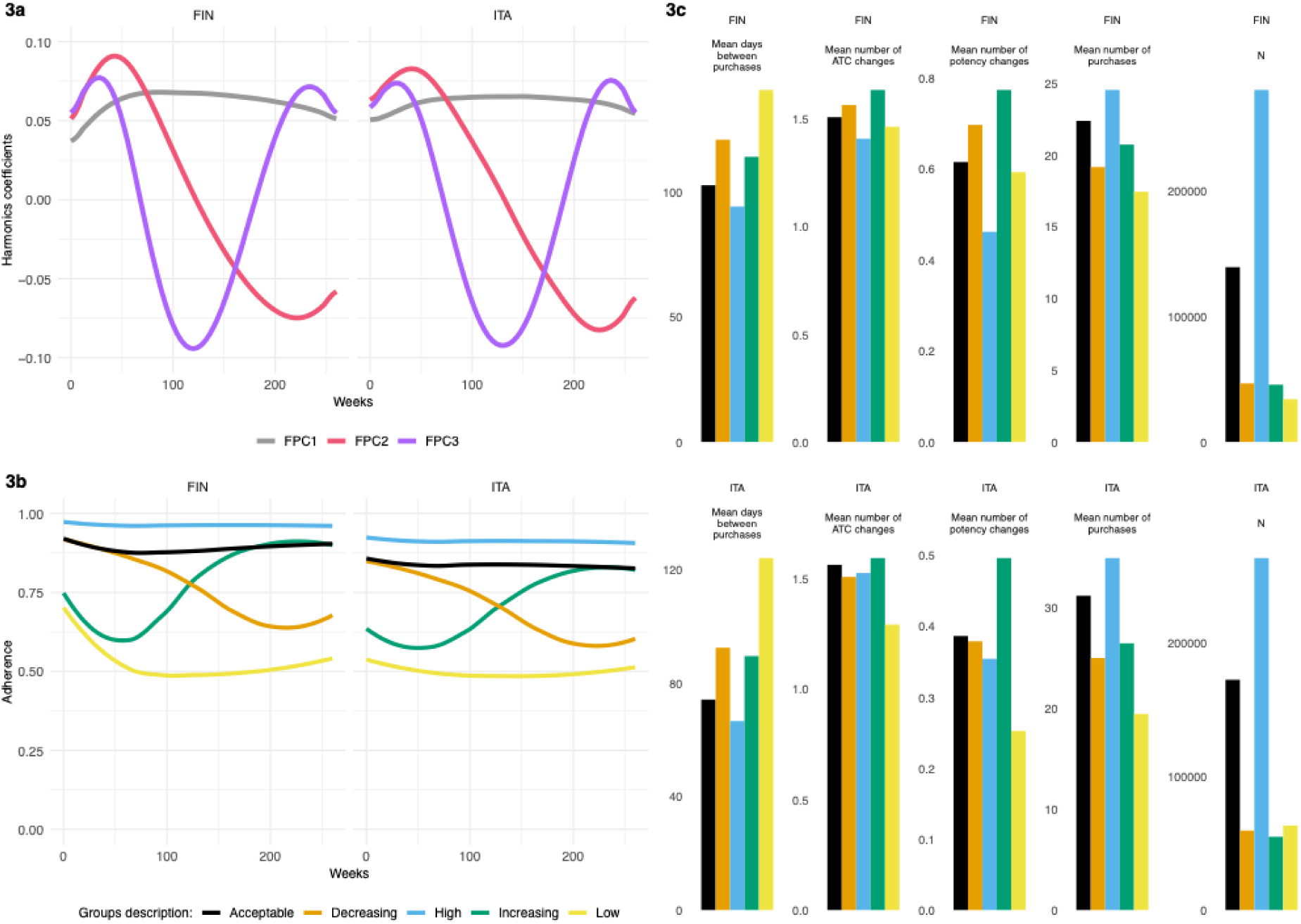
Latent trajectories clustering (a) First three Functional Principal Components; (b) Medioids of the six groups identified by the clustering; (c) Summary statistics for each of the groups (mean days between purchase, mean number of ATC changes, mean number of changes of potency, mean number of purchases, numerosity)

### Trajectories clustering reveals overall good statin taking behaviour

Five distinct adherence trajectory groups were identified in both countries. Clustering individuals using k-means on the first three FPCA scores identified five trajectory groups that together explained more than 75% of the total variance; median trajectories are shown in Figure 3b. These groups comprised high adherence (Finland: 51.36%; Italy: 42.95%), acceptable adherence (Finland: 25.50%; Italy: 28.11%), decreasing adherence (Finland: 8.54%; Italy: 9.69%), increasing adherence (Finland: 8.35%; Italy: 8.94%), and low adherence (Finland: 6.24%; Italy: 10.31%). Further group-level characteristics are reported in Supplementary Data 4. Compared with threshold-based adherence classifications, trajectory clustering provided a more granular representation of longitudinal medication-taking behaviour by explicitly capturing adherence dynamics (Supplementary Methods).

### Determinants of drug adherence

Baseline demographic, socioeconomic, and clinical factors were associated with adherence decline. To examine correlates of declining adherence, we defined a binary outcome distinguishing individuals with declining trajectories from those with stable optimal adherence (Equation 6). Predictors were derived exclusively from information available prior to the start of the five-year trajectories. We evaluated 131 predictors in Finland and 112 in Italy, spanning health conditions, medication use, socioeconomic and demographic characteristics, and outpatient care indicators (see Predictors in Materials section).

We fitted logistic regression, Lasso regression, and Light-GBM models to assess associations with declining adherence. Results from logistic regression are presented for interpretability; corresponding results from the Lasso and Light-GBM models are reported in the Supplementary Results (Supplementary Data 7–10). Logistic regression estimates are reported as odds ratios with false discovery rate–adjusted p-values (Figure 4; Supplementary Data 5 and 6).

**Figure 4:**
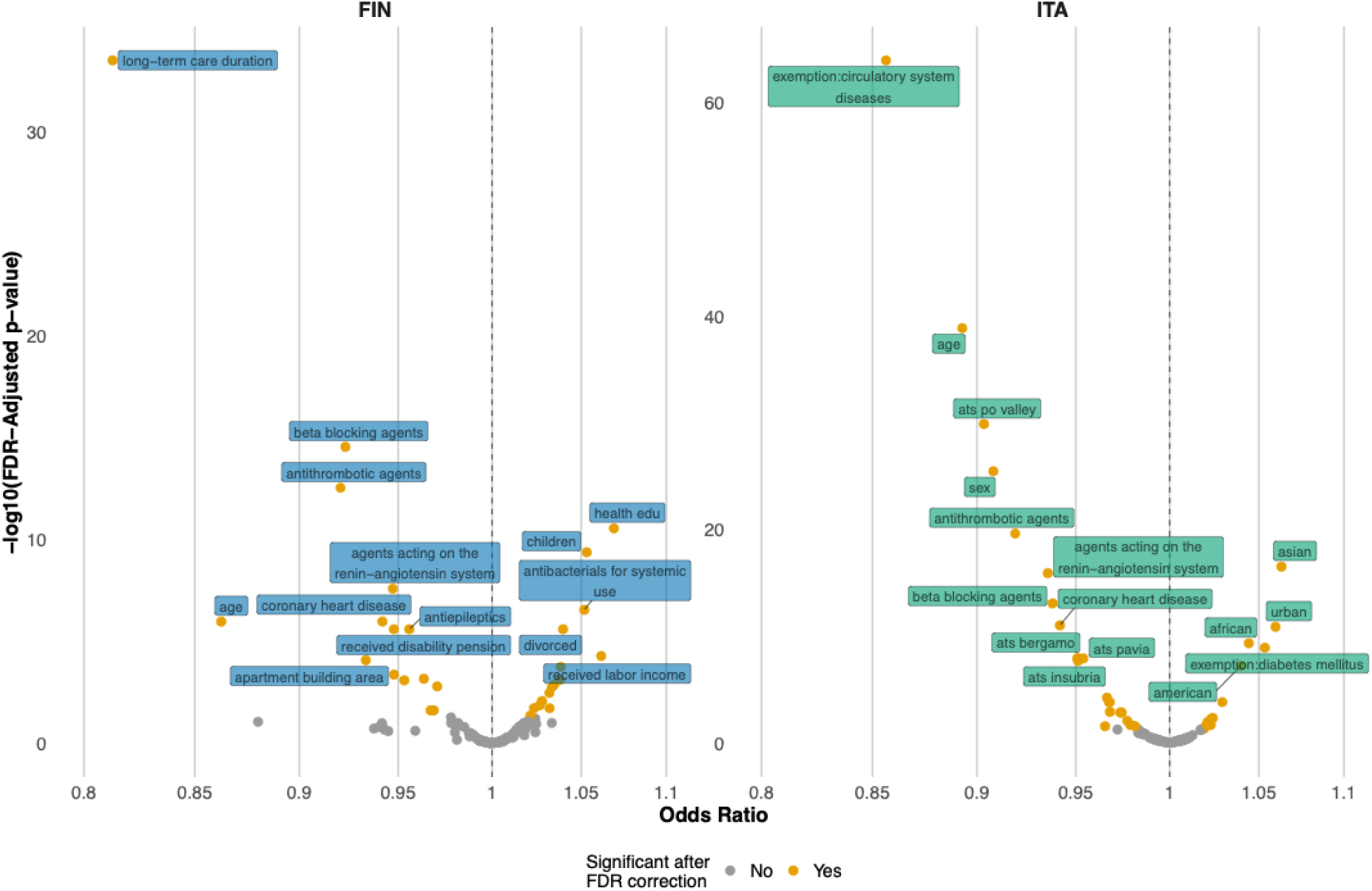
Declining adherence determinants Volcano plot of the odds ratio from the logistic model, adjusted for false discovery rate

### Country-specific associations

Associations between predictors and declining adherence differed by national context. In Finland, longer duration in long-term care was associated with lower odds of declining adherence (OR per SD increase 0.81, 95% CI 0.79–0.83), as was older age (OR 0.86, 95% CI 0.82–0.91). Higher odds of decline were observed among individuals with children and those who were divorced. An educational background in a health-related discipline and several economic indicators, including receipt of labour income or social allowances, were also associated with increased odds of decline. Clinically, diagnoses of type 2 diabetes, coronary heart disease, stroke, and gallstones were each associated with reduced odds of declining adherence.

### Medication-wide associations

Several medication classes showed consistent associations across cohorts. Using ATC codes, medication histories were compared directly between countries (Figure 5). In both Finland and Italy, antithrombotic agents, renin–angiotensin system agents, beta-blockers, and antidiabetic medications were associated with lower odds of declining adherence. Other associations differed between cohorts. Nervous system medications showed contrasting patterns, with psycholeptics associated with increased odds of decline in Finland but decreased odds in Italy. Additional cohort-specific associations were observed for gastrointestinal, anti-inflammatory, antibacterial, cardiac, and antihypertensive medications.

**Figure 5:**
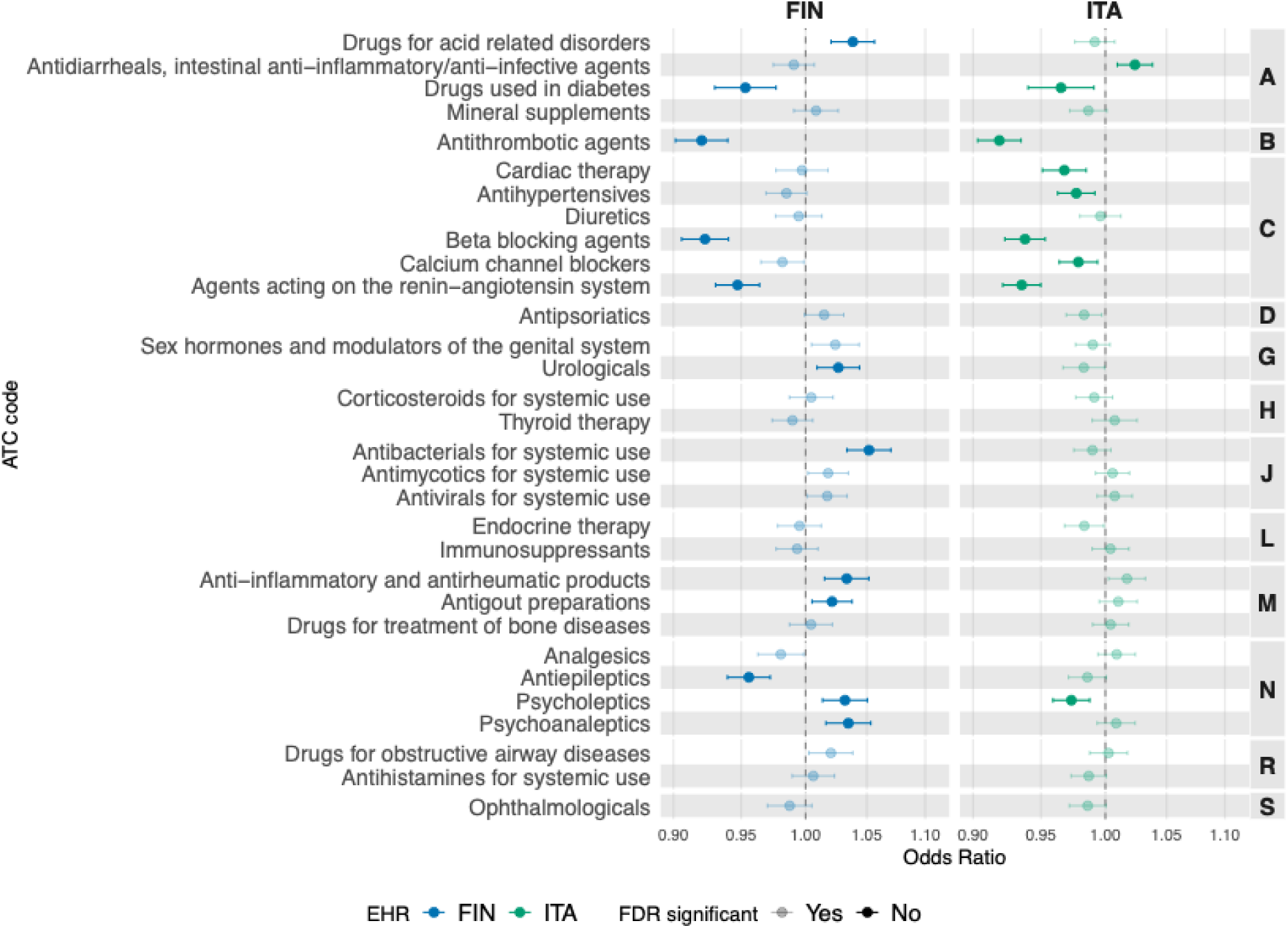
Treatments-wide comparison Treatments-wide comparison, odds ratios with 95% confidence interval corrected for false discovery rate

Baseline predictors showed limited ability to identify declining trajectories. Prediction models demonstrated modest performance in both cohorts. In Finland, AUC values ranged from 59.86% to 60.62% across models, while in Italy they ranged from 61.53% to 62.28%. Neither dimensionality reduction nor modelling non-linear relationships led to substantial improvements in predictive performance (Supplementary Table 3; Supplementary Figures 6 and 7).

## Discussion

Using functional data analysis, we identified distinct longitudinal patterns of adherence to statins, revealing notable similarities and variations between two national populations, despite differences in healthcare organisation and prescribing practices. The overall average adherence was relatively high in both cohorts, consistent with other population-based studies^20,21^, although adherence was modestly higher in Finland. These differences likely reflect variations in healthcare policies^22^, prescribing practices^23^, and broader sociocultural contexts that shape medication-taking behaviour.

Across both countries, three functional components captured most of the variation in adherence trajectories. The dominant pattern, explaining approximately 60% of the variance, reflected stable adherence over time, indicating that most individuals maintained consistent statin use once treatment was established. The remaining components captured increasing, decreasing, and non-monotonic adherence patterns, highlighting variability in long-term behaviour among a non-trivial subset of patients. Clustering based on these components identified five robust adherence subgroups consistent in both cohorts. While the high-adherence group was the largest in both countries, low and declining adherence trajectories were more prevalent in Italy^24^, suggesting systematic differences in adherence maintenance across settings.

Differences in prescribing patterns further illustrate the role of healthcare context. Italian patients were more frequently treated with high-potency statins and experienced more frequent potency switching than Finnish patients^25,26^. These differences may reflect variation in national guidelines, clinical risk assessment, or prescribing norms, and suggest that adherence trajectories emerge from interactions between individual behaviour and healthcare system characteristics. Importantly, the presence of substantial low and declining adherence groups in both countries indicates that adherence challenges remain widespread, even in settings with high overall statin use.

Medication- and disease-related factors showed consistent associations with adherence trajectories across cohorts. Drug classes typically prescribed for cardiometabolic conditions and secondary prevention were associated with greater adherence persistence, supporting the notion that perceived disease severity and treatment salience contribute to sustained medication use. By contrast, associations involving nervous system medications differed between countries, pointing to heterogeneity in how mental health and neurological conditions intersect with long-term medication behaviour. These differences may reflect variation in care structures, stigma, or continuity of mental health services across healthcare systems.

Socioeconomic and demographic factors were strongly associated with declining adherence^13,27,28^. Older age was consistently associated with more stable adherence, whereas economic disadvantage was associated with higher odds of decline in both countries. Sex differences were context dependent: no meaningful differences were observed in Finland^29^, whereas males in Italy were more likely to exhibit declining adherence, suggesting that gendered health behaviours may vary across sociocultural settings. Indicators of social support also played a role. In Finland, being divorced or having children was associated with declining adherence, whereas participation in long-term care services was associated with more stable trajectories, potentially reflecting increased supervision and care coordination.

Geographic context further shaped adherence patterns. In Finland, residence in sparsely populated northern regions was associated with declining adherence, suggesting barriers related to service access. In Italy, urban residence, particularly in the Milan area, was associated with higher odds of declining adherence, whereas less urbanised regions showed more stable patterns, possibly reflecting differences in healthcare burden or continuity of care. Unexpectedly, a health-related educational background was associated with declining adherence in Finland, highlighting the complexity of behavioural factors and suggesting that health knowledge alone may not translate into sustained medication use.

This study has several strengths. Modelling adherence as a continuous, time-evolving behaviour using FDA allowed us to characterise longitudinal adherence dynamics without relying on arbitrary thresholds and to identify latent adherence trajectories that are not captured by conventional measures. The cross-national design, drawing on population-level data from Finland and Italy, enabled the examination of both shared and context-specific determinants of adherence across distinct healthcare systems. In addition, the comprehensive inclusion of demographic, socioeconomic, clinical, and medication-related predictors, combined with the use of both interpretable statistical models and machine-learning approaches, strengthened the robustness of the findings by allowing linear and non-linear associations to be assessed consistently across cohorts. Limitations should also be acknowledged. Adherence trajectories were modelled without explicitly accounting for time-varying clinical events during follow-up, such as disease progression or the onset of new comorbidities, which may influence medication-taking behaviour and alter adherence patterns, even though they should theoretically influence the adherence upward more than towards a decline. Capturing such dynamics would require alternative longitudinal modelling strategies and represents an important direction for future work. Despite the breadth of baseline information considered, predictive performance for identifying declining adherence remained modest, suggesting that adherence trajectories are shaped by complex behavioural and contextual factors that are not fully captured in routinely collected health records.

Overall, these findings indicate that long-term statin adherence reflects an interplay between individual behaviour, clinical context, and healthcare system characteristics. While precise prediction of declining adherence remains challenging, the identification of reproducible adherence trajectories and shared determinants across populations may inform population-level strategies aimed at supporting sustained medication use.

## Material and Methods

### Lombardy region administrative data (LR)

Lombardy is the most populated region in Italy (10 million inhabitants). Healthcare and administrative data are recorded for the whole population; in this case, we have information about all individuals over 30 years of age on 1st January 2020 registered with the public healthcare system. The dataset includes almost 7 million individuals (n=7,335,190) and covers January 2012 to June 2023. The data resource comprises diverse healthcare information from regional electronic health records, encompassing inpatient diagnosis, outpatient visits, state financial exemptions, demographics, vaccinations and medications§.

### Lombardy pharmacy drug register

The regional pharmaceutical and in-hospital delivered pharmaceutical registers include information about reimbursable prescribed medication purchased in territorial and hospital pharmacies. We evaluated only purchases from 2015 onward to have a minimal evaluation window and ensure completeness. Each row encodes information about the date, quantity, AIC code (identification code for medicinal products for human use in Italy, from which the number of units per package and dosages can be identified), and ATC code.

### FinRegistry (FINR)

The FinRegistry^30^ dataset includes all individuals alive in 2010 in Finland and all their relatives, for more than 7 million people (n=7,166,416). For consistency in the available data, we restrict the population target to only the indexed individuals, hence those who were alive in Finland in 2010, which is a total of 5 million people. Finregistry encompasses several population registries that record a vast spectrum of socioeconomic, geographic, and health information.

### Kela drug purchase register

The Social Insurance Institution of Finland (Kela) is a governmental agency that provides basic financial security for all residents in Finland. The Kela drug purchase register contains records of reimbursable prescription medications purchased from pharmacies in Finland from 1995 to 2021. Each entry includes details such as the purchase date, the medication’s ATC code, the number of packages bought, and the Nordic Article Number (VNR), which provides information about the medication’s quantity and dosage.

### Cohort definition - LR

Starting from the Lombardy pharmacy drug register, we excluded all purchases from 2012 to 2014 to ensure the consistency of the registry. We selected all individuals with at least five years of continued statins purchases from the first purchase recorded and at least five purchases in the five years, therefore excluding early treatment discontinuation. We also excluded individuals who discontinued the treatment defined as those with a gap between purchases greater than one year plus the number of previous units bought, for example if the previous purchase was of 28 tablets, discontinuation was defined as a gap with the following purchase of 365 + 28 days.

### Cohort definition - FINR

Starting from the Kela drug purchase register, we excluded all purchases from 1995 to 1997 to ensure the consistency of the registry. We selected all individuals with at least five years of continued statins purchases from the first purchase recorded and at least five purchases in the five years, excluding individuals who discontinued the treatment (defined as those with a gap between purchases greater than one year plus the number of previous units bought).

### Predictors –LR

Predictors for the RL cohort were extracted from several datasets; hereafter they are described singularly with the respective variables defined:

### Pharmacy drug register

The medication predictors were retrieved from the Lombardy pharmacy drug register. For each three-digit ATC code (101 different strings), a dichotomous variable was encoded (excluding lipid modifying agents “C10”); if at least one purchase was recorded in the previous three years from the start date, it was given a value of one; otherwise, zero.

### Inpatient data

One primary diagnosis and five secondary diagnoses were encoded in ICD9CM^31^ for each hospitalisation. The diagnosis and ATC codes from the pharmaceutical registry were combined to define twenty-two diseases. We also defined a measure of overall health with the Multisource Comorbidity Score as defined by Corrao, G. *et al.* ^32^.

### Outpatient data

The outpatient care visit dataset records only the general speciality of the visit. The predictors were encoded as dichotomous variables by the type of speciality visit recorded (one for at least one, zero otherwise), except for laboratory visits, where the total number of visits was used as a continuous variable since the distribution was not as heavily zero-inflated.

### Demographic data

Age in years, sex, country of birth (encoded in continents), and ATS (”Agenzia di Tutela della Salute”, the eight local healthcare provider branches) were retrieved from the personal information dataset. We also defined a variable with the degree of urbanisation, the municipality of domicile (one for urban, zero for otherwise), based on the Italian Institute of Statistics (ISTAT)^33^.

### Medical cost exemptions registry

This registry includes all medical cost exemptions provided by the region. They are encoded in broad categories of reason: economic (with different income levels), disability, age or disease-specific (e.g. diabetic patients within a range of income are exempt from paying for diabetes medications). We considered all exemptions released in the previous three years before the start of the treatment. The definitions of the exemption codes are reported in Lombardy’s official documentation^34^. We added three broader categories of variables representing any exemption for financial, invalidity and disability reasons.

### Vaccination registry

Each vaccination and the type of vaccine delivered are recorded in the registry. We defined one variable for each vaccination type and included a variable specifying the number of vaccinations delivered. We applied the three years before the start cutoff also to the vaccination data.

### Predictors – FINR

All diseases, medication and socioeconomic information were extracted following the FinRegistry matrices pipelines^35^. We selected a subset of 128 endpoints from the core endpoints defined by the FinnGen^36^ phenotype. Furthermore, it included the Charlson comorbidity index^37^. We defined the dichotomous variables for the medications using the first three digits of ATC codes (101, which is the same as RL in this case). We selected a subset of 198 socioeconomic variables extracted from multiple registries^38^ and described in detail by Hartonen et al.^39^.

### Adherence

Adherence is defined as the Medication Possession Ratio ^16^, calculated at each subsequent purchase, excluding the first purchase. If a change of statin (ATC code) happened, we restarted the adherence calculation, excluding the purchase when the change occurred. We set a limit for each adherence measured to one. This rule is defined to remove the effect of the stockpiling behaviour on the outcome.

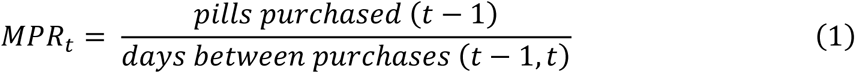

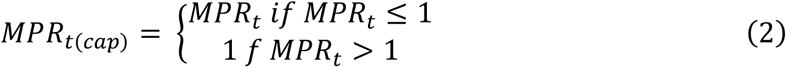

To calculate (2), we extracted the number of tablets for each package from AIC (RL) and VNR (FINR) codes and multiplied them by the number of packages purchased. Finally, we used a definition of statin potency based on the dosage and the type of statin^40^.

### Functional data analysis

For each individual, we performed smoothing of 𝑀𝑃𝑅_𝑡(𝑐𝑎𝑝)_ on the days of treatment using penalised splines ^41^. The rationale for using penalised splines is that they allow us to capture non-linear trends in the data while avoiding overfitting to short-term fluctuations. In practice, we represent each trajectory 𝑦(𝑡) as a linear combination of spline basis functions,

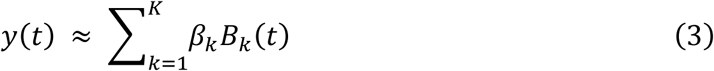

where 𝐵_𝑘_(𝑡) are piecewise polynomial basis functions defined over a set of knots (in our case, corresponding to every observed time point, hence different for each individual). A penalty term is added to the least-squares loss to control the smoothness of the curve, in the form:

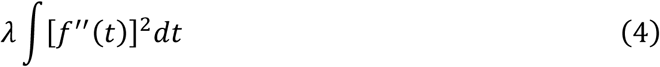

where 𝑓^′′^(𝑡) is the second derivative of the fitted curve, and 𝜆 is the smoothing parameter. Larger values of 𝜆 penalise curvature more heavily, leading to smoother curves. We manually selected a single smoothing parameter 𝜆 that achieved a balance between smoothness and fidelity to the observed data and could be applied consistently across all individuals, given the high computational cost of choosing an optimal 𝜆 for each individual through cross-validation. The distribution of the generalised cross-validation error is reported in Supplementary Figure 1 to show the overall goodness of fit of the chosen 𝜆.

When smoothing, we retained the first measurement that exceeded the five-year observation window to anchor the spline fit to an actual observation. The smoothed curves were evaluated on a standardised grid covering the same five-year period, with time points every four weeks. Finally, we re-expressed the smoothed curves using an unpenalised spline basis with knots placed at four-week intervals, ensuring that all individuals’ functions were represented in a homogeneous basis for subsequent analysis.

We performed Functional Principal Component Analysis^19^ on the representation with homogeneous basis. We calculated the Functional Principal Components Scores, allowing us to rewrite each individual’s curve as a summation of a mean curve plus a weighted set of independent principal curves, each of them representing a pattern:

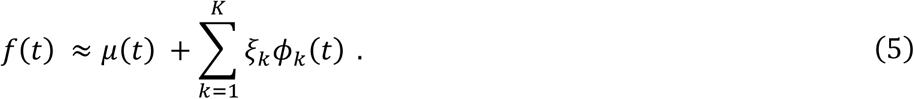

Where 𝑓(𝑡) is the observed functional data or curve measured (in our case, the smoothed adherence measurement across days); 𝜇(𝑡) is the average function or mean curve across the entire population. It captures the central trend or pattern common to all subjects; 𝜉_𝑘_ are the Functional Principal Component Scores: these scalar scores quantify the magnitude and direction of how much an individual’s functional data differs from the mean curve in the direction of the k^th^ eigenfunction. They reflect individual-specific variations or deviations; 𝜙_𝑘_ are the Eigenfunctions or Functional Principal Components), these functions represent distinct modes of variation derived from the covariance structure of the data.

We clustered the individuals based on the first three component scores through the k-means algorithm^42^. We choose the number of clusters to retain based on the percentage of variance explained by the groups (at least 75%).

### Unsupervised predictors selection

To reduce sparsity and correlation between dichotomous variables, we removed low-frequency categories (< 0.01) and variables with lower frequency from clusters of highly associated variables. To cluster variables, we defined distances between each pair within a subset separately (e.g. distances across medications, distances across diseases) with the complement to one of the phi coefficients^43^ (also known as the Matthews Correlation Coefficient or Yule phi coefficient) for dichotomous variables.

Then, we performed hierarchical clustering with average linkage^44,45^ and clustered at 0.4 distance. Hence, variables with an association higher than 0.6 were retained in the same cluster; we kept only the variable with the highest frequency from each cluster. We also removed highly correlated continuous variables from the socioeconomic selection based on a pairwise correlation higher than 0.7.

### Prediction models

Building on the functional clustering, we defined a binary outcome to discriminate optimal from declining adherence. For individual *i*:

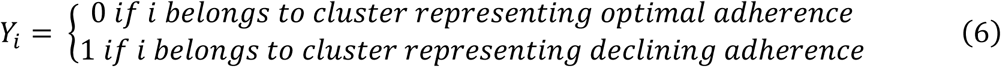

Since we are mainly interested in capturing the determinants of declining adherence, we only retained the individuals belonging to these two groups in the analysis of determinants, therefore comparing individuals with declining adherence and those with stable high adherence. Missing values of the predictors (mainly on the municipality of residence and related fields) were handled by imputation with predictive mean matching^46^ for continuous variables; for categorical variables, a new variable was encoded, representing the missing value if needed. We split both datasets into training (70%) and testing (30%) and under-sampled the controls (optimal adherence group) in the training phase to obtain a balanced set.

To evaluate the determinants of declining adherence, we fitted three different models: standard logistic regression, LASSO -penalised logistic regression^47^, and Light-GBM^48^ gradient-boosted trees. We choose Light-GBM because of its ability to handle sparser data with many hot-encoded variables. All models were fitted with tidymodels^49^ pipelines and evaluated on AUC (Area Under Curve); also, accuracy, sensitivity and specificity were reported Supplementary Results Table 3.

#### Logistic regression

We fitted one model, comprehensive of all selected predictors specified in terms of the *ith* statistical unit as:

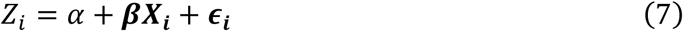

Where 𝑍_𝑖_ is the logit of the binary outcome 𝑌_𝑖_ defined in (4); 𝑋_𝑖_ is the vector of predictors. Parameters coefficient estimates, standard errors, p-values and 0.95 confidence intervals were extracted. P-values were adjusted with False Discovery Rate (Benjamini–Hochberg procedure^50^) to account for multiple testing.

#### Lasso regression

We fitted a LASSO regression to evaluate whether further parameter selection could improve prediction:

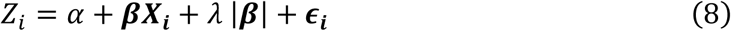

Where 𝑍_𝑖_ is the logit of the binary outcome 𝑌_𝑖_ defined in (4); 𝑋_𝑖_ is the vector of predictors; 𝜆 is the shrinkage parameter for 𝜷, the value of 𝜆 was tuned with 10-fold cross-validation. The criteria for selecting 𝜆 were to be the most regularising 𝜆 within one standard deviation of the 𝜆 with the minimum error.

#### Light-Gradient boosting

We fitted a more complex machine learning algorithm to understand if it could improve prediction. We fitted a Light-Gradient boosting machine^48^, a tree-based model. Compared with other tree-based models, Light-GBM reduces the computational burden with large datasets in the training phase and suits many binary features. The model’s hyperparameters were optimised for AUC in ten-fold cross-validation on a grid search.

## Data availability

Data dictionaries for FinRegistry are publicly available on the FinRegistry website (www.finregistry.fi/finnish-registry-data). Access to the FinRegistry data can be obtained by submitting a data permit application for individual-level data to the Finnish social and health data permit authority, Findata (https://asiointi.findata.fi/). The application includes information on the purpose of data use; the requested data, including the variables, definitions of the target and control groups, and external datasets to be combined with FinRegistry data; the dates of the data needed; and a data utilization plan. The requests are evaluated case by case. Once approved, the data are sent to a secure computing environment (Kapseli) and can be accessed within the European Economic Area and within countries with an adequacy decision from the European Commission. Data availability for Lombardy region data is publicly available on the Epidemiological Observatory of the Lombardy Region website (www.osservatorioepidemiologico.regione.lombardia.it/wps/portal/site/osservatorio-epidemiologico/DettaglioRedazionale/collaborazioni-con-gli-enti/daas+2-0/red-daas-2-0). Access to the Lombardy region data can be obtained by submitting a project application for individual-level data. The application includes information on the purpose of data use; the requested data, including the variables, definitions of the target and control groups; the dates of the data needed; and a data utilization plan. The requests are evaluated case by case. Once approved, the data are sent to a secure computing environment (Daas 2.0).

## Contributions

Study design: A.C, A.G., F.I. Data analysis: A.C., M.F., and K.M.L.. Results interpretation: A.C., K.M.L., M.F., M.P., E.D.A., A.G., F.I.. Writing original draft: A.C.. All authors were involved in further drafts of the manuscript and revised it critically for content.

## Acknowledgments

We are deeply grateful to all individuals in the Finnish and Lombardy Region, whose data made this study possible. I would also like to thank the entire *THL*, *FinRegistry*, *ARIA*, and *Osservatorio Empidemiologico Lombardo* teams for making the data available for the study. The present research is part of the activities of “Dipartimento di Eccellenza 2023-2027” (Department of Mathematics), Polytechnic of Milan. We would also like to acknowledge Luisa Zuccolo and Claudia Giambartolemi for their suggestions on the draft of the manuscript.

## Code availability

The analysis code used to produce the results is available on GitHub at: https://github.com/ht-diva/Drug-adherence-trajectories.

