## Supplementary Information for "Longitudinal patterns and determinants of statin adherence in over one million individuals from Finland and Italy"

#### Supplementary methods

##### Clustering comparison with binary adherence

We evaluated the trajectories clusters comparing their discrimination capacity compared to the cross sectional estimation of adherence as a binary trait (threshold of 0.8) in the Italian cohort. We selected six variables, two demographic: age (AGE) and sex (SEX); two health-related: previous diagnosis of coronary heart disease (CHD) and previous diagnosis diabetes (DIAB); and two adherenc related: number of purchases (PURCH) and mean lag between purchases (LAG). For each of the variables we computed the weighted variance within groups (1) for the continous variables (AGE, PURCH and LAG) and the Shannon entropy (2) for the binary ones (SEX, CHD, DIAB). We compared these variances among the two clustering (binary vs trajectories groups) with bootstrapping (1000 bootstrap samples) computing the empirical p-value and 95% confidence intervals with quantiles of the bootstrap samples (3).

$$V_w = \frac{1}{N} \sum_{i \in c} (x_i - \bar{x_c})^2 \cdot n_c \tag{1}$$

$$H_w = \frac{1}{N} \sum_{c \in C} n_c \, (- \sum_{v \in \{0,1\}} p_{c,v} \cdot \ln(p_{c,v}) \tag{2}$$

$\boldsymbol{Input}$:

$X = (x_i, \dots, x_N)$ $variable\ of\ interest$

$C1 = (c_i^1, \dots, c_N^2)$ $binary\ clustering\ labels$

$C2 = (c_i^2, \dots, c_N^2)$ $trajectories\ clustering\ labels$

$B = 1000$ $bootstrap\ iterations$

$\boldsymbol{Step\ 1}$: $observed\ difference$

$v1 = V_w\ or\ H_w(X, C1);$

$v2 = V_w\ or\ H_w(X, C2);$

$\Delta = v1 - v2$

$\boldsymbol{Step\ 2}$: $boostrap$

$\boldsymbol{for}\ b = 1\ to\ B\ \boldsymbol{do}$

$X_b = sample(X)$

$v1_b = V_w\ or\ H_w(X_b, C1_b)$

$v2_b = V_w\ or\ H_w(X_b, C2_b)$

$\Delta_b = v1_b - v2_b$

$\boldsymbol{Step\ 3}$: $inference$

$$p.value\ bootstrap = \frac{1}{B}\sum_{b}\mathbf{I}(|\Delta_b - \Delta| \geq |\Delta - 0|)$$
$$ci\ 95\%\ bootstrap = (\Delta_{B,0.025};\ \Delta_{B,0.975}) \quad (3)$$

### Supplementary results

#### Comparison with binary adherence

We report separate tables for continuos traits and binary traits. Since the continuos variables were scaled the variance within groups can be interpreted as the percentage of variance explained by the groups (the total variance is one). The trajectories clustering with improved the AGE variance explained by the groups of very little (0.2%) while sensibly more the LAG and PURCH (10% and 6%), all bootstrap p-values equal to zero. For the binary variables we see a significant but small difference in SEX and CHD (0.0012 and 0.0021) and a non-significant one for DIAB. This analysis shows how statistically the trajectories clustering offer a better stratification and representation then the binary classification of the drug taking behaviour.

| Variable | Observed difference | Bootstrap p-value | 95% CI lower | 95% CI upper | Variance binary | Variance groups |
|---|---|---|---|---|---|---|
| AGE | 0.0019 | 0.0000 | 0.0016 | 0.0022 | 0.9982 | 0.9963 |
| LAG | 0.1031 | 0.0000 | 0.1011 | 0.1053 | 0.6850 | 0.5819 |
| PURCH | 0.0614 | 0.0000 | 0.0603 | 0.0628 | 0.7000 | 0.6386 |

*Table 1: Continuos tratits tests results*

| Variable | Observed difference | Bootstrap p-value | 95% CI lower | 95% CI upper | Entropy binary | Entropy groups |
|---|---|---|---|---|---|---|
| SEX | 0.0012 | 0.0000 | 0.0010 | 0.0014 | 0.9940 | 0.9928 |
| CHD | 0.0021 | 0.0000 | 0.0018 | 0.0024 | 0.5557 | 0.5536 |
| DIAB | 0.0000 | 0.5310 | 0.0000 | 0.0001 | 0.7017 | 0.7017 |

*Table 2: Binary tratits tests results*

#### Detailed logistic OR analysis

**Finland**

Time spent in the long-term care program (a predictor from the The Care Register for Social Welfare representing the total number of treatment days by institutional care and residential services of social welfare, which can be interpreted as a measure of social support) is strongly associated with reduced odds of adherence decline. Specifically, each standard deviation increases in care duration corresponded to an odds ratio of 0.81 (95% CI: 0.79–0.83), indicating that individuals receiving extended long-term care are less likely to experience a drop in adherence. Similarly, older age is associated with a stable pattern (OR = 0.86, 95% CI: 0.82–0.91), suggesting that adherence improves slightly with increasing age. While having children (OR = 1.05, 95% CI: 1.03–1.07) and being divorced (OR = 1.03, 95% CI: 1.02-1.05) are associated with higher odds of adherence decline. An educational background in a health discipline was linked to adherence decline (OR = 1.07, 95% CI: 1.05–1.09). Among economic indicators, receiving labour income was associated with increased odds of declining adherence (OR = 1.06, 95% CI: 1.03–1.09), as was receiving any income support (OR = 1.04, 95% CI: 1.02–1.06) and receiving other social allowances (OR = 1.03, 95% CI: 1.01–1.04). Conversely, living in the Lapland region (postcode starting with 9) was associated with higher odds of declining adherence (OR = 1.03, 95% CI: 1.01–1.05). From a clinical perspective, diagnosis of type 2 diabetes (OR = 0.97, 95% CI: 0.94–0.99), coronary heart disease (OR = 0.94, 95% CI: 0.92–0.97), stroke (OR = 0.96; 95% CI: 0.94-0.98) and gallstones (OR = 0.97, 95% CI: 0.95–0.99) were all associated with a lower risk of declining adherence.

**Italy**

Age was inversely associated with adherence decline (OR = 0.89, 95% CI: 0.88–0.90), suggesting that older individuals were less likely to experience a drop in adherence. Sex was also a significant factor, with females showing reduced odds of declining adherence compared to males (OR = 0.91, 95% CI: 0.89–0.93). Ethnic background also influenced adherence; individuals born in Asia (OR = 1.06, 95% CI: 1.04–1.07), Africa (OR = 1.04, 95% CI: 1.03–1.06), and the Americas (OR = 1.04, 95% CI: 1.02–1.06) were significantly more likely to experience a decline in adherence. Clinically, a diagnosis of coronary heart disease (CHD) was associated with higher adherence (OR = 0.94, 95% CI: 0.93–0.96).

Among medical and financial exemptions (economic relief issued by the state to specific population categories based on income and diseases diagnosis), exemption for any circulatory system disease (ES_002) was linked to a decrease in the odds of declining adherence (OR = 0.86, 95% CI: 0.84-0.87), while exemption for diabetes mellitus (ES_013; OR = 1.05, 95% CI: 1.04–1.07) and hypertension (ES_031; OR = 1.02, 95% CI: 1.02–1.04) were linked to an increase in the odds of declining adherence. Other exemptions, such as for unemployment with low income (ES_E02, OR = 1.03, 95% CI: 1.01–1.04) and social allowance recipients (ES_E03, OR = 1.02, 95% CI: 1.01–1.03), were associated with worse adherence, as well as receipt of a minimum pension (ES_E04; OR = 1.02, 95% CI: 1.1-1.4) was associated with adherence decline.

Geographic and sociocultural context played a role in the Italian cohort. Individuals classified as residing in urban areas had significantly higher odds of adherence decline (OR = 1.06, 95% CI: 1.04–1.08). All regional health administration zones (ATS) also showed a negative association with declining adherence when compared to the most urban zone (Milan): for instance, citizens followed by the ATS Val Padana (south-east part of Lombardy) have lower odds of declining adherence (OR = 0.90, 95% CI: 0.88-0.92) while the other ATS have OR close to 0.95. Lastly, individuals who visited the emergency room (OR = 1.03, 95% CI: 1.01–1.05) and those who underwent plastic surgery (OR = 0.98, 95% CI: 0.96–0.99) exhibited distinct risk profiles, with emergency room attendance being a factor associated with declining adherence and plastic surgery associated with the regular trajectory.

We can directly compare information between countries from medications purchases, thanks to international coding systems (ATC). From the analysis of the odds ratios in Figure 5, similarities and discrepancies arise. For example, antithrombotic agents (B01, FIN: OR = 0.92, 95% CI: 0.90-0.94, ITA: OR = 0.92, 95% CI: 0.90–0.93), agents acting on the renin-angiotensin system (C09, FIN: OR =0.94, 95% CI: 0.93-0.96, ITA: OR = 0.93, 95% CI: 0.92–0.93), beta-blockers (C07, FIN: OR = 0.92, 95% CI: 0.90-0.94, ITA: OR = 0.94, 95% CI: 0.92–0.95), and antidiabetics (A10, FIN: OR = 0.95, 95% CI = 0.93-0.98, ITA: OR = 0.96, 95% CI:0.94-0.99) were associated with reduced odds of belonging to a decreasing adherence group in both cohorts (OR=0.92 in Finland and Italy). Differences emerged in the nervous system medications (chapter N), we observed that antiepileptics (N03, OR=0.95, 95% CI: 0.93-0.97) and psychoanaleptics (N06, OR = 1.03, 95% CI: 1.02-1.05) were significantly associated with adherence

in Finland, while psycholeptics (N05) showed contrasting associations across cohorts: positively associated with adherence decline in Finland (OR = 1.03, 95% CI: 1.01-1.05) and negatively in Italy (OR=0.97, 95% CI: 0.96-0.99). In Finland, drugs for acid-related disorders (A02, OR = 1.04, 95% CI: 1.02.1.06), anti-inflammatory and antirheumatic products (M01, OR = 1.03, 95% CI: 1.02-1.05), and antibacterials (J01; OR = 1.05, 95% CI: 1.03-1.07) were positively associated with declining adherence (whereas no significant association was detected in Italy. Conversely, antidiarrheals and intestinal anti-inflammatory agents (A07, OR = 1.02, 95% CI: 1.01–1.04) were linked to increased odds of decreased adherence in Italy but not Finland. Finally, cardiac therapy agents (C01, OR = 0.97, 95% CI: 0.95-0.98) and antihypertensives (C02, OR = 0.97, 95% CI: 0.96-0.99) were linked to lower odds of declining adherence only in Italy.

#### Lasso features analysis

A few patterns stand out in Supplementary Figure 3. Some factors have broadly similar associations in both cohorts (e.g., markers of cardiovascular disease and related therapies, disability pensions, and exemptions), while others differ in size or even direction, suggesting context-specific effects. Geographic and demographic variables (such as rural domicile, age, language background, or region/ATS area) show clear signals, and multiple concomitant medication classes (e.g., antithrombotics, beta-blockers, agents acting on the renin–angiotensin system, antiepileptics) also line up consistently also in LASSO model. It stands out that for sex we see a large effect in the Italian cohort, while for the Finnish one it is shrinked to zero, denoting no differences.

#### Light-GBM features analysis

Feature importance was assessed based on three metrics: Gain (contribution to model accuracy), Cover (number of observations impacted), and Frequency (number of times the variable was used in tree splits). We report the most important points by Gain (Suppelemary Figure 4).

**Finland**

The Light-GBM model found age and year of birth to be the most dominant predictors in the model (gain of 14.6% and 8.5%, respectively), while long-term care duration emerged as the third most influential variable (gain = 8.4%). Among clinical and medication features, antithrombotics (B01, gain = 7.2%), beta-blockers (C07, gain = 5.3%), renin-angiotensin system agents (C09, gain = 2.5%), antidiabetics (A10, gain = 2.3%), coronary heart disease, and antibacterials (J01, gain = 1.4%) featured prominently. The Light-GBM model also highlighted the importance of secondary prevention in determining adherence through coronary heart disease (gain = 2.2 %) and comorbidities through the Charlson comorbidity score (gain = 1.3%), which captures the overall disease burden.

Socioeconomic variables such as received labour income (gain = 3.1%), pension (gain = 7.2%), total benefits (gain = 2%), and total income (gain = 1.3%) contributed to the model. Variables like education in health fields (gain = 1.9%) and non-Finnish/Swedish mother tongue (gain = 1.7%) retained modest but consistent importance. Region-level factors, including inter-municipal migration (gain = 1.3%) and permanent resident fraction (gain = 2.3%), also contributed to the model.

**Italy**

On the Light-GBM side, the most important feature in gain is antithrombotic agents (B01, gain = 14.5 %), which account for the highest contribution to the model's predictive performance, followed by coronary heart disease diagnosis (gain = 10.2%) and age (gain = 9.8%). Exemption for circulatory conditions (ES_002; gain = 5.6%) and beta-blockers (C07, gain = 4.9%) also showed high importance. Sex (gain = 3.3%) and cardiac therapies (C01, gain = 3.2%) also contributed to the prediction.

Other variables such as renin-angiotensin system agents (C09, gain = 2.2%), Multisource Comorbidity Score (MCS; gain = 2.2%), and stroke (gain = 1.9%) rank moderately high, indicating their relevance in specific patient subgroups. Acid-related disorder medications (A02, gain = 1.9%) and calcium channel blockers (C08, gain = 1.6%) are notable contributors to pharmacologic features. Urban classification also appears in the importance ranking (gain = 1.2%). Among outpatient visits, cardiology (gain = 3.4%) and laboratory tests (1.8%) are the visits that contributed the most. Finally, the exemption for low-income and rare diseases is among the bigger contributors (gain = 1.6%).

#### Models' performances

| | Finland | | | | Italy | | | |
|---|---|---|---|---|---|---|---|---|
| **MODEL** | **AUC** | **ACC** | **SENS** | **SPEC** | **AUC** | **ACC** | **SENS** | **SPEC** |
| LOGISTIC | 0.601 | 0.572 | 0.573 | 0.571 | 0.616 | 0.566 | 0.555 | 0.614 |
| LASSO | 0.599 | 0.569 | 0.568 | 0.573 | 0.615 | 0.562 | 0.548 | 0.620 |
| LGBM | 0.606 | 0.595 | 0.603 | 0.547 | 0.623 | 0.574 | 0.566 | 0.611 |

*Table 3: Prediction performances of logistic regression (LOGISTIC), LASSO regression (LASSO), and Light-Gradient boosting machine (LGBM)*

### Supplementary Figures

##### Supplementary Figure 1

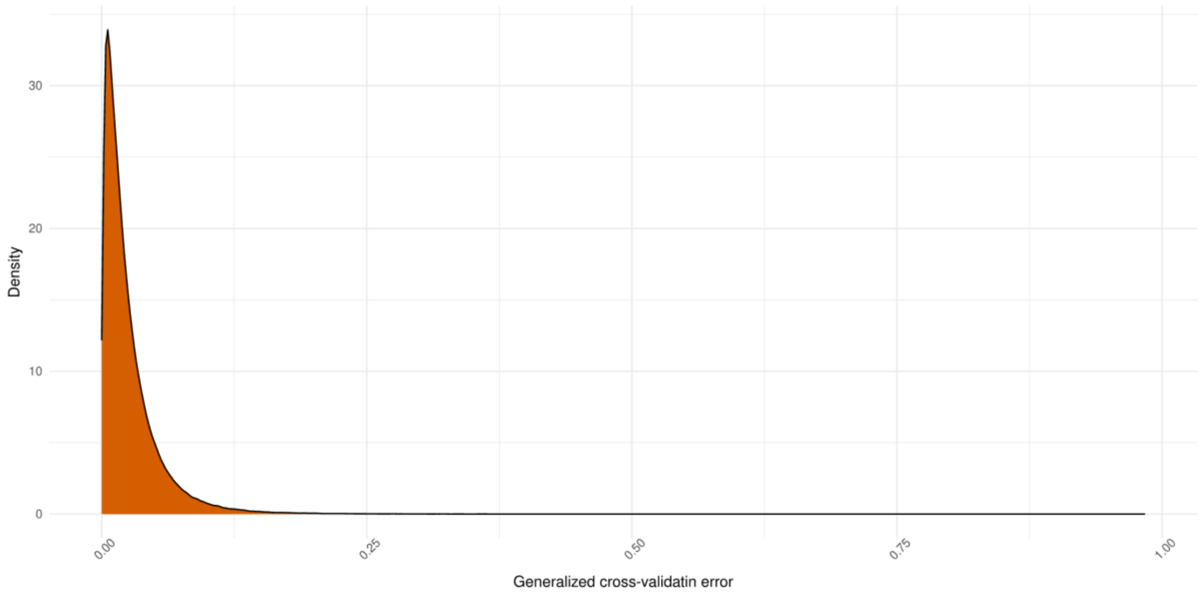


*Figure 1: Distribution of generalized cross-validation error for the reconstruction of the adherence trajectories*

#### Supplementary Figure 2

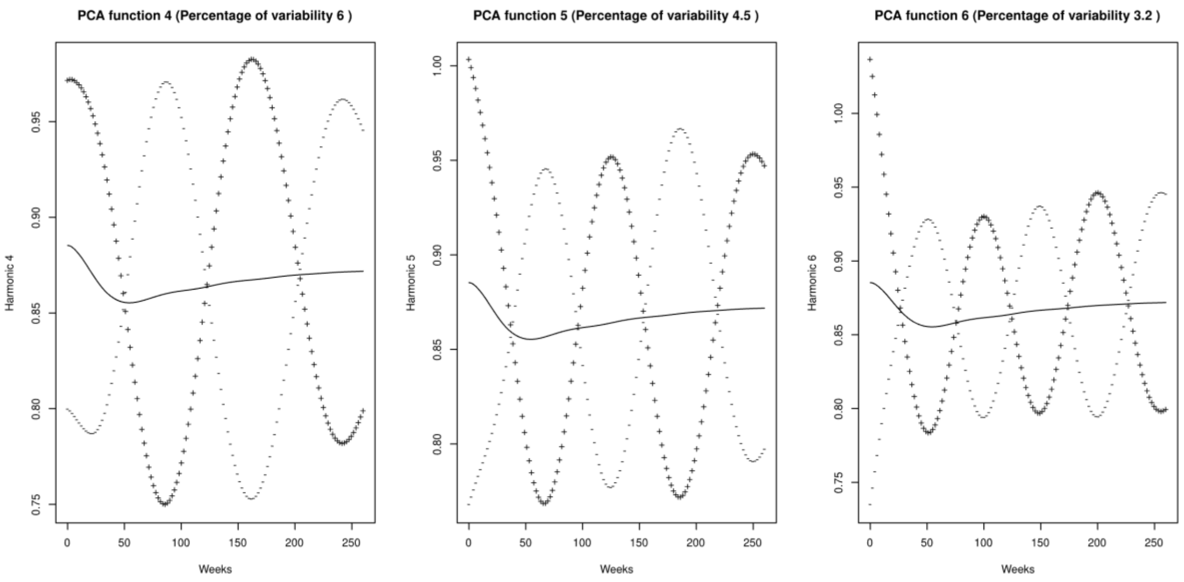


*Figure 2: 4th to 6th FPC harmonics of adherence trajectories of the Finnish cohort*

#### Supplementary Figure 3

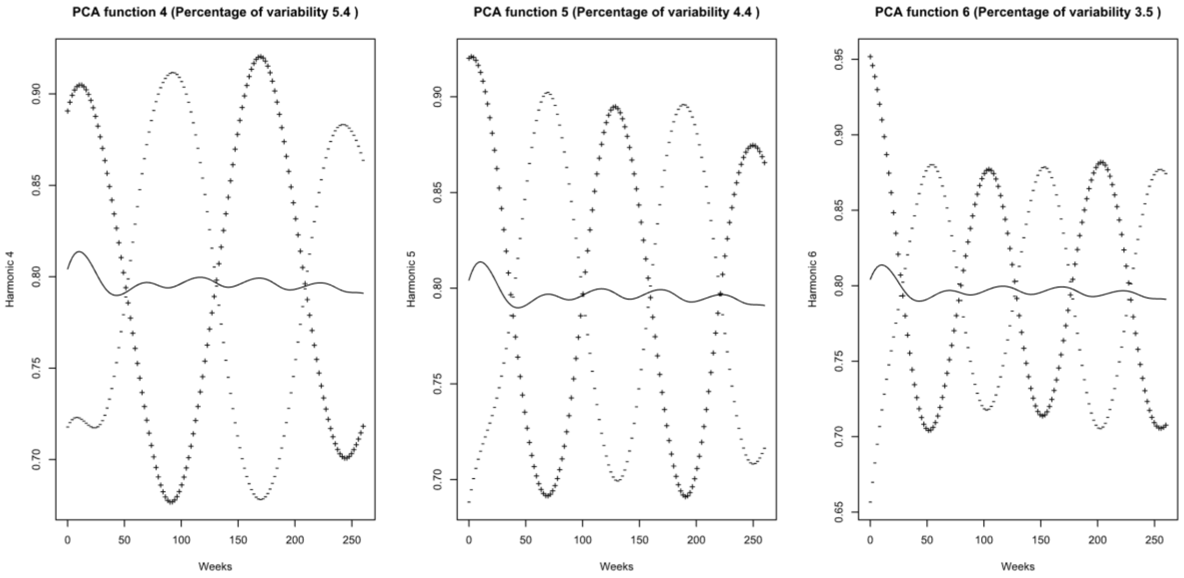


*Figure 3: 4th to 6th FPC harmonics of adherence trajectories of the Italian cohort*

#### Supplementary Figure 4

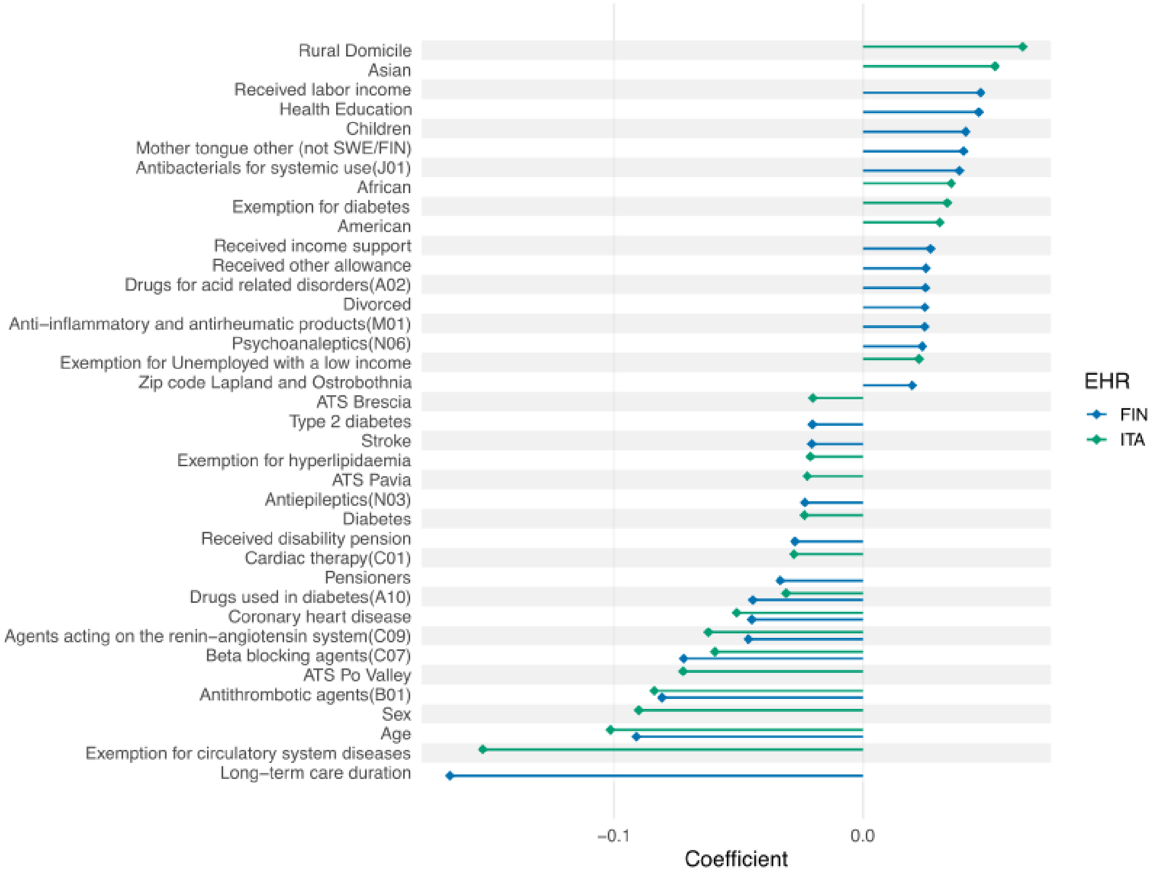


*Figure 4: Non-zero coefficients from the LASSO regression combined (whenever possible) between Italy (green) and Finland (blue)*

#### Supplementary Figure 5

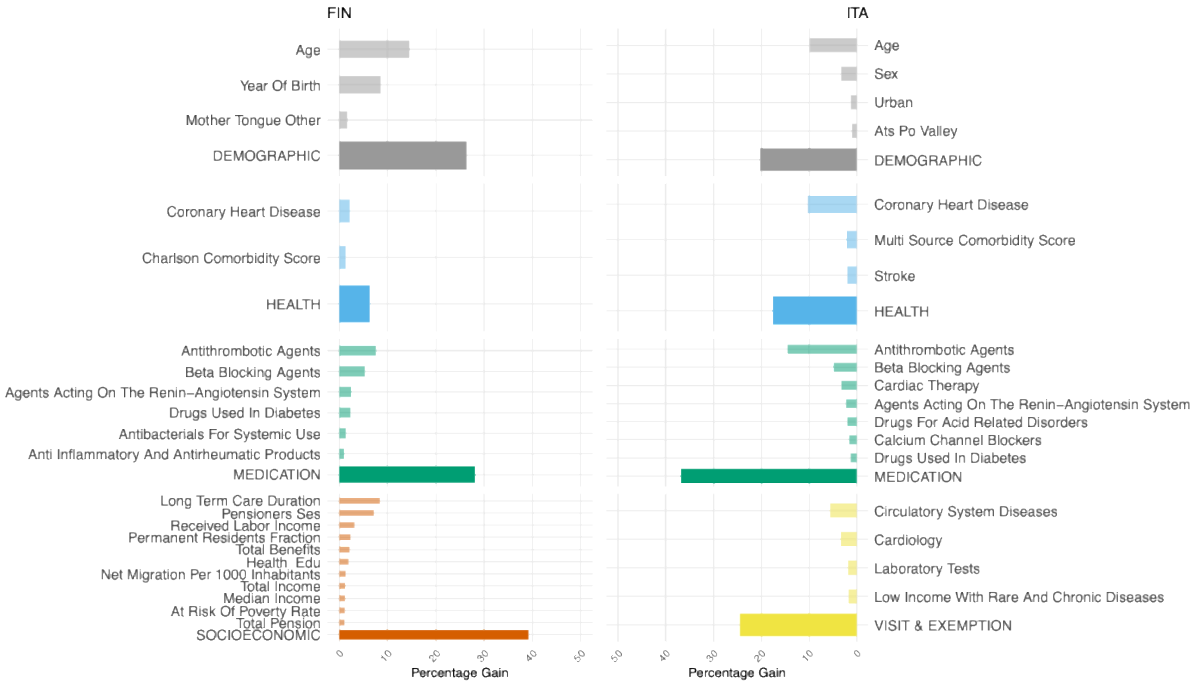


*Figure 5: Light-GBM importance in terms of gain grouped by macro-category and the macro-category aggregate gain*

#### Supplementary Figure 6

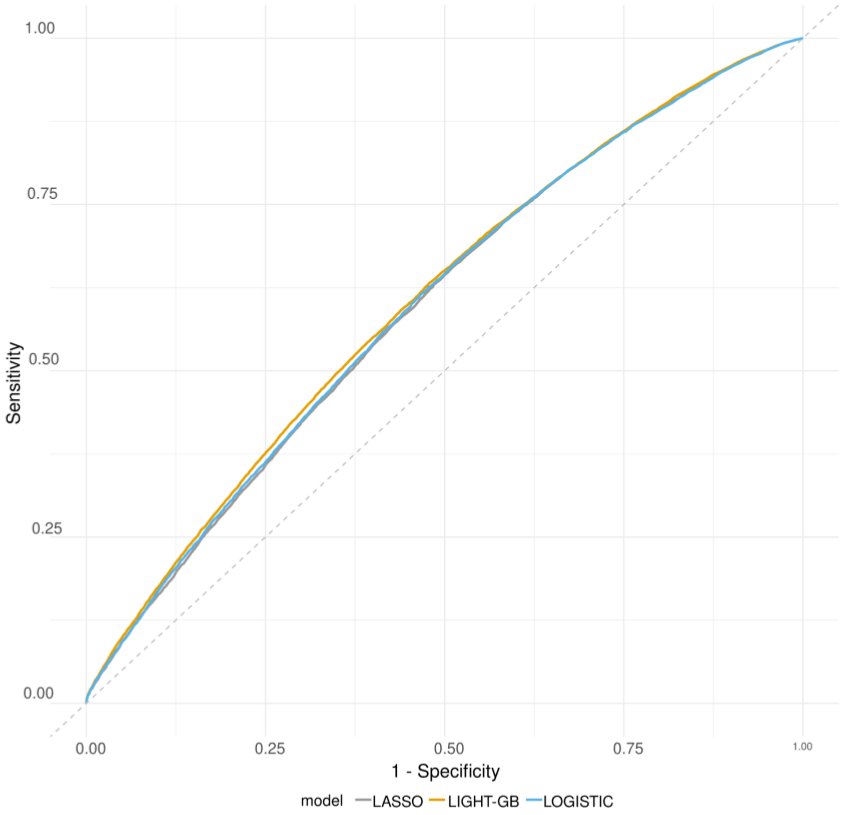


*Figure 6:ROC curves of the three models (logistic, lasso and LGBM) of the Finnish cohort*

#### Supplementary Figure 7

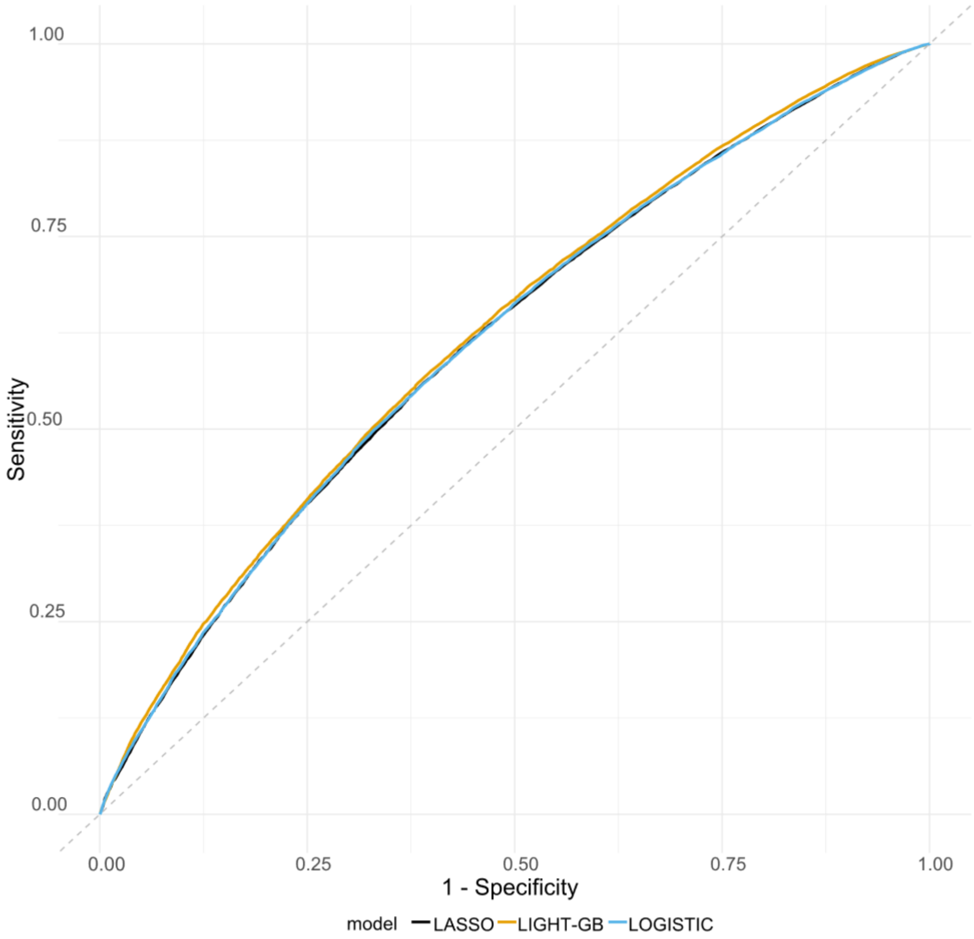


*Figure 7: ROC curves of the three models (logistic, lasso and LGBM) of the Italian cohort*
